# Understanding the association between mental health knowledge and mental health service utilization among Black adults

**DOI:** 10.1101/2021.11.06.21266012

**Authors:** Aderonke Bamgbose Pederson, Alexander C. Tsai, Devan Hawkins, Judith T. Moskowitz, Lisa Dixon

## Abstract

Mental health knowledge limitations may contribute to the treatment gap among Black adults. We conducted an online cross-sectional study of Black adults in the United States (n = 262, aged 18-65 years) from diverse ethnic backgrounds (African-Americans, African immigrants, Afro-Caribbean immigrants). Gamma regression using generalized linear models was used to estimate the associations between mental health knowledge and willingness to seek help from mental health professionals. After adjusting for age, education and ethnicity, participants with higher specific knowledge about mental health (such as recognition of schizophrenia as a mental illness) were 26% more likely to report willingness to seek help from a mental health professional for personal and emotional problems (RR = 1.26, CI: 1.12 – 1.41, p < 0.001). Knowledge building interventions (such as psychoeducation) that seek to increase specific knowledge (rather than general knowledge) may correlate more strongly with utilization of mental health services among Black adults.

## Introduction

In the US, Black adults represent 12% of the United States population and comprise nearly 20% of those affected by mental illnesses (1, 2). At the same time, they have lower mental health service utilization (25%) compared to white adults (37.6%) (3), which may lead to more severe illness and worse overall mental health outcomes. Black adults are also grossly underrepresented in clinical research (4). Black adults thus experience substantial mental health disparities (5-9).

Andersen’s behavioral model to understand health service utilization has been used extensively in research investigating factors that optimize health service engagement (10). It considers three main factors: predisposing characteristics (demographics, social structures, and health beliefs or knowledge); enabling resources (personal, family and community context); and need (perceived and evaluated) (11). Research over the last half a century has refined and expounded on this model (10-14). We focused on knowledge as a predisposing factor in the use of mental health services in a sample of Black adults. The benefits of health knowledge and education around physical illnesses are well accepted; however, the benefits of *mental* health knowledge are not as widely recognized within health systems and the community (5, 6). Importantly, Andersen specified the importance of general versus specific knowledge and suggested that limitations in understanding how knowledge influences utilization of mental health services in much of the existing (early) research was due to absence of these distinctions.

Research on psychoeducation provides some context for the value of knowledge in driving service utilization. Psychoeducation is defined as any education that educates patients or families (by increasing knowledge) about a mental illness in order to improve mental health outcomes (15, 16). Several studies show that psychoeducation builds knowledge but may not lead to a change in behavior (e.g., use of mental health services) (16). Previous studies have generated mixed findings, and there remain several open questions about the nature of the association between different forms of knowledge (general and specific) and mental health service utilization (10, 11, 13). Knowledge is malleable and represents an area of focus that can potentially be used to address low utilization of mental health services among Black adults in a way that is low cost with a high potential for effective broad dissemination and implementation (17, 18).

A potentially important related factor is that Black adults have greater stigma toward mental illness compared with white adults (1, 19, 20). Addressing mental illness stigma involves improving mental health knowledge and utilization of mental health services (7-9). Stigma is often influenced by cultural beliefs and values (7-9). Stigma related to mental illness refers to negative knowledge, attitudes and beliefs held by individuals or the community; stigma is a fundamental cause of health inequities and leads to further discriminatory experiences in mental health care, which is of particular significance for Black adults (21-26). Previous research suggests that mental illness stigma in Black adults may lead to delay in initiation of treatment, poor adherence to treatment and, consequently, increased morbidity and mortality (1, 19, 20, 27). Knowledge is a primary component of the stigma framework and is the most common target of stigma-reducing interventions (27, 28); therefore, we examined the association between mental health knowledge and help seeking behavior and how stigma interacts with mental health knowledge.

The aim of our study was to assess specific areas of mental health knowledge (general mental health knowledge and recognition of specific mental health conditions) and their associations with willingness to seek help (for personal and emotional problems and for suicidal ideation) from a mental health professional. We also assessed whether anticipated behavior reflecting stigma towards individuals living with a mental illness moderated the association between mental health knowledge and willingness to seek help from a mental health professional. We hypothesized that greater knowledge would be associated with greater willingness to use mental health services and that the association between mental health knowledge and likelihood of seeking help for mental health problems would be weaker for those with higher stigma compared to lower stigma. This study will contribute to understanding the association between mental health knowledge and its effectiveness in improving willingness to engage in mental health care for Black adults.

## Methods

### Overview

In September-October 2020, we conducted an online cross-sectional survey among African-Americans, African immigrants, and Afro-Caribbean immigrants (n=262) in the United States. The convenience sample of participants was recruited using email and social media (Twitter and Facebook) in partnership with local community-based organizations (World Relief Chicago, the United African Organization, Refugee One) who shared the survey with their members. Eligibility was restricted to people who: 1) identified as Black, African-American, African or Afro-Caribbean; 2) were 18-65 years of age; 3) were residing in the United States at the time of the study; and 4) were English speaking. Participants completed informed consent procedures prior to study activities using the online consent forms and received $25 compensation for their time. Data collection occurred after obtaining approval by the Institutional Review Board at Northwestern University (IRB ID #: STU00213136).

### Assessments

We collected socio-demographic information including age, gender, race, ethnicity, marital status, education, employment status, income and insurance status.

### The Mental Health Knowledge Schedule (MAKS) (29)

The MAKS is composed of six questions about stigma-related mental health knowledge (knowledge about help-seeking, recognition, support, employment, treatment and recovery) and six questions designed to elicit whether the respondent can accurately identify common conditions as mental illnesses or not (depression, stress, schizophrenia, bipolar disorder, substance use disorders, grief). Each of the 12 items is scored on a 5-point Likert scale (ranging from “strongly disagree” to “strongly agree”); higher total scores represent greater knowledge. In our analysis, we summed the first six questions to calculate a general knowledge score (that could range from 6-30), which was then analyzed as a single variable. The other six questions about accurately identifying mental illnesses were analyzed as separate variables: respondents were categorized as being able to accurately identify mental illnesses if they strongly agreed that depression, schizophrenia, bipolar disorder and substance use disorder were mental illnesses and/or if they strongly agreed that stress and grief were not mental illnesses. In population-based studies, the MAKS has demonstrated moderate to substantial test-retest and internal consistency reliability (30). In our study sample, the Cronbach’s alpha for the knowledge score was 0.42.

### The General Help-Seeking Questionnaire (GHSQ) (31)

The GHSQ was designed to assess intentions to seek help from 11 different sources (e.g., partner, friend, or helpline) for two different mental health concerns (personal/emotional problems, suicidal ideation). Each item is scored on a 7-point Likert-type scale (ranging from “extremely unlikely” to “extremely likely”). For this study, we focused specifically on the two items that assessed intentions to seek help (for personal/emotional problems or for suicidal ideation) from mental health professionals such as counselors or psychologists.

### Reported and Intended Behavior Scale (RIBS) (25)

The RIBS measures the reported social distance from people with a mental health problem in the past, currently, and in the future. In this study we reported only the measure of stigma behavior in the future. The RIBS measures intended future behavior using four questions, each scored on a 5-point Likert-type scale (agree strongly to disagree strongly, with an option to answer don’t know). Each question is phrased similarly to the following: “In the future, I would be willing to [live with/work with/be a neighbor to/have a close relationship with] someone with a mental health problem.” The 4 items are summed to generate a total future intended behavior score that ranges from 4-20. In the original scale development studies, the RIBS subscale on intended behavior showed good test-retest and internal consistency reliability (25). In our study sample, the Cronbach’s alpha was 0.67.

### Data Analysis

Summary data was calculated for the following variables age group, gender, marital status, education, income, insurance, ethnicity, time in the US, and citizenship status. The percentage of respondents who believed that different conditions were mental illnesses was also calculated. Additionally, the distribution of the likelihood of seeking help for a mental health problem from a mental health professional was examined.

In order to estimate the associations between mental health knowledge and willingness to seek help from mental health professionals, Gamma regression using generalized linear models in SAS Version 9.3 (SAS Institute Inc., Cary, NC, USA) was used. We specified two dependent variables: willingness to seek help from a mental health professional for a personal/emotional problem, and willingness to seek help from a mental health professional for suicidal thoughts. We fitted a series of gamma regression models alternately specifying the general knowledge score and the 6 variables accurately identifying conditions as mental illnesses as the explanatory variable of interest. For the relationship between the total mental health knowledge score and willingness outcome, the rate ratio (RR) represents the average change in willingness per unit change in the total mental health knowledge score. For the questions assessing study participants’ beliefs about different conditions being mental illnesses, the RR represents how many times higher the willingness score is for those who believe the condition is a mental illness compared with those who do not. Univariable and multivariable-adjusted models (including age, education, and ethnicity as covariates) were fitted to the data. Thus we fitted a total of 28 regression models (7 unadjusted and 7 adjusted estimates for each of 2 outcomes).

We assessed future intended stigma behavior as a moderator of the relationship between mental health knowledge and willingness to seek mental health services. First we divided the sample into those with lower stigma (based on median score of 12 or less) and higher stigma scores (based on median score of greater than 12) according to their intended future stigma behavior (based on the RIBS scale). Then fitted the same regression models described above. Additionally, we fitted a regression model to the pooled sample that contained a product term between the low/high stigma variable and the knowledge questions to assess for a potential interaction.

## Results

Table 1 shows the demographic and sample characteristics of the participants. We also report the mean willingness to seek mental health care from a mental health professional.

**Table 1.**
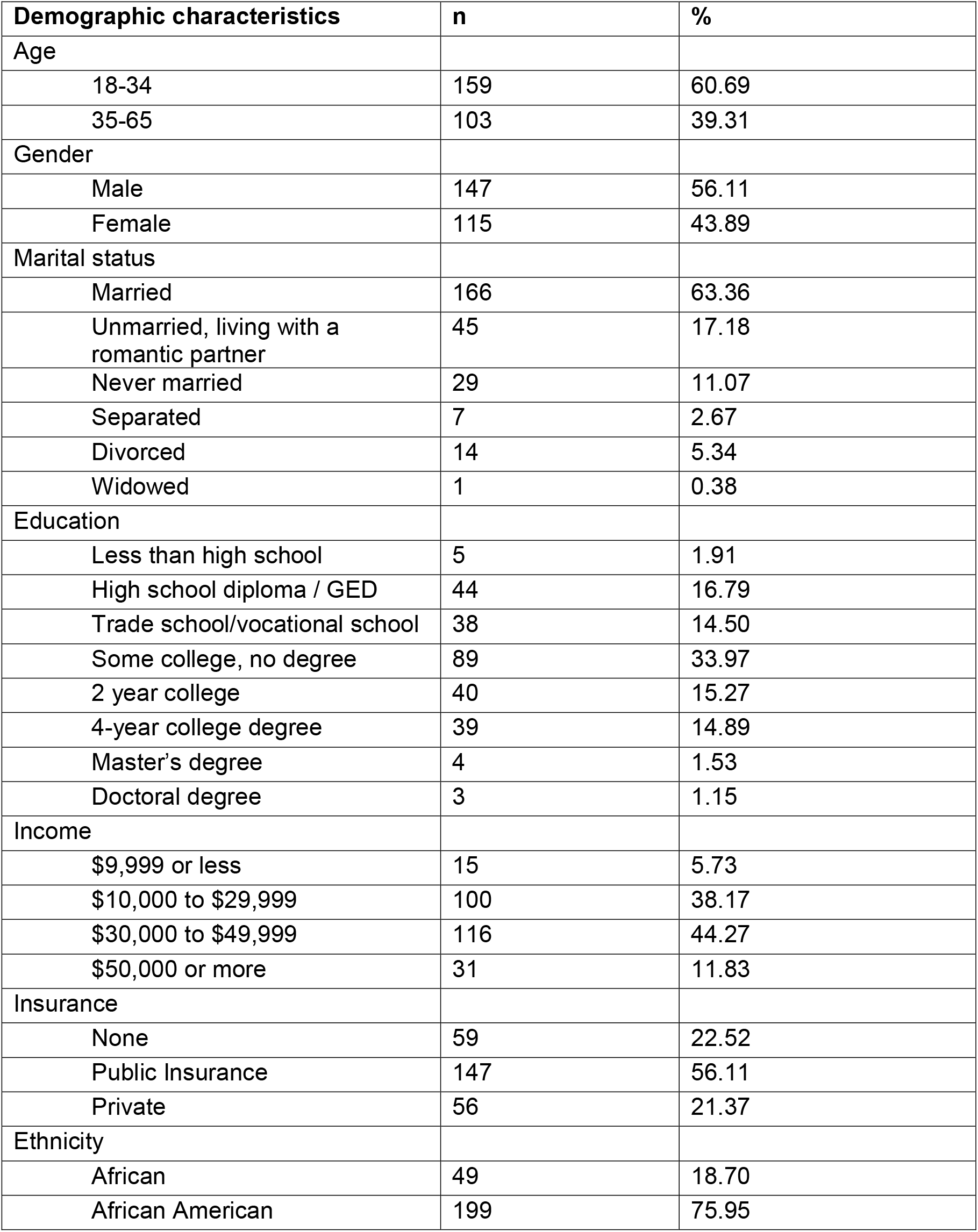
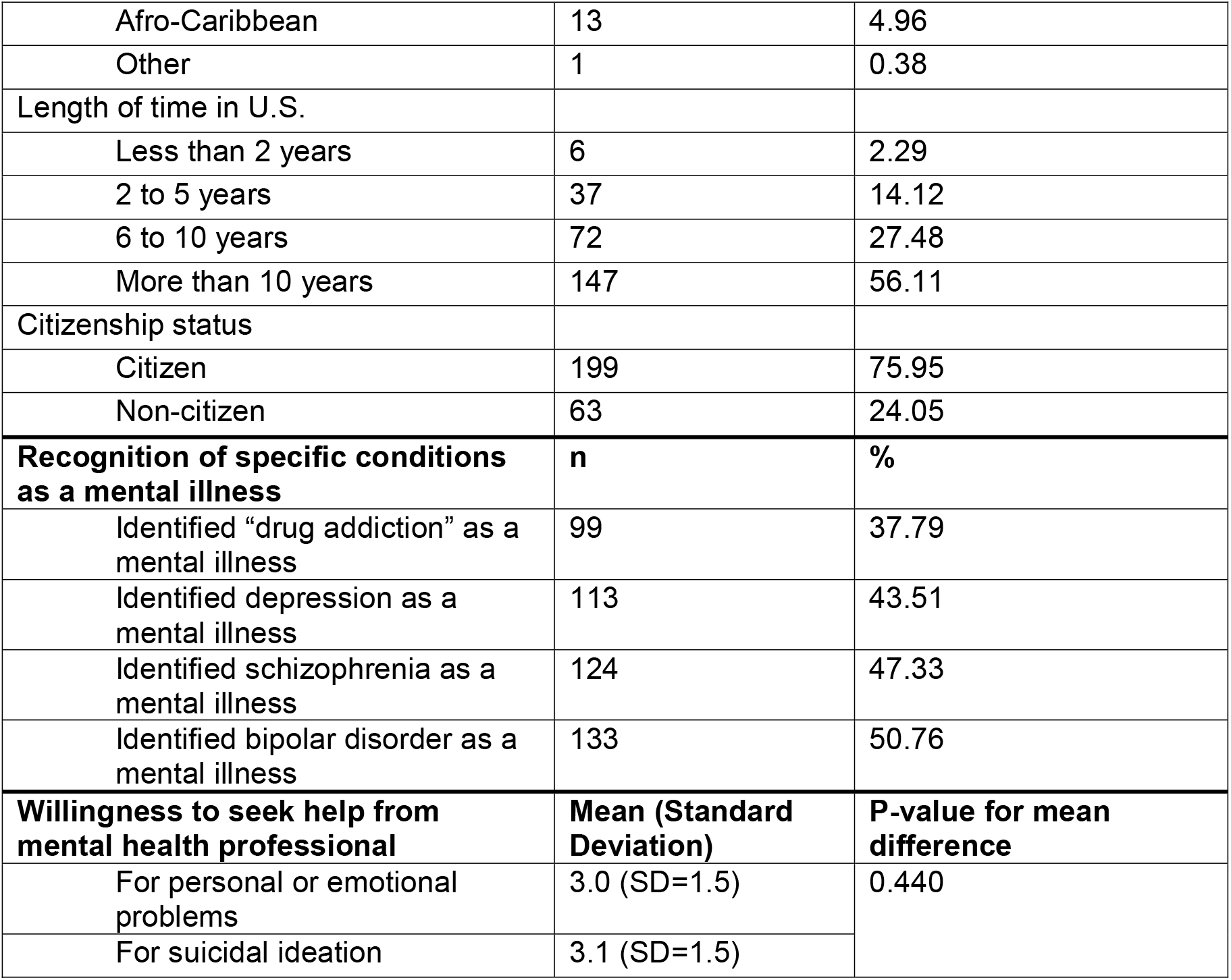
Descriptive demographics and sample characteristics.

| **Demographic characteristics** | **n** | **%** |
| --- | --- | --- |
| Age |  |  |
| 18-34 | 159 | 60.69 |
| 35-65 | 103 | 39.31 |
| Gender |  |  |
| Male | 147 | 56.11 |
| Female | 115 | 43.89 |
| Marital status |  |  |
| Married | 166 | 63.36 |
| Unmarried, living with a romantic partner | 45 | 17.18 |
| Never married | 29 | 11.07 |
| Separated | 7 | 2.67 |
| Divorced | 14 | 5.34 |
| Widowed | 1 | 0.38 |
| Education |  |  |
| Less than high school | 5 | 1.91 |
| High school diploma / GED | 44 | 16.79 |
| Trade school/vocational school | 38 | 14.50 |
| Some college, no degree | 89 | 33.97 |
| 2 year college | 40 | 15.27 |
| 4-year college degree | 39 | 14.89 |
| Master's degree | 4 | 1.53 |
| Doctoral degree | 3 | 1.15 |
| Income |  |  |
| \$9,999 or less | 15 | 5.73 |
| \$10,000 to \$29,999 | 100 | 38.17 |
| \$30,000 to \$49,999 | 116 | 44.27 |
| \$50,000 or more | 31 | 11.83 |
| Insurance |  |  |
| None | 59 | 22.52 |
| Public Insurance | 147 | 56.11 |
| Private | 56 | 21.37 |
| Ethnicity |  |  |
| African | 49 | 18.70 |
| African American | 199 | 75.95 |
| Afro-Caribbean | 13 | 4.96 |
| Other | 1 | 0.38 |
| Length of time in U.S. |  |  |
| Less than 2 years | 6 | 2.29 |
| 2 to 5 years | 37 | 14.12 |
| 6 to 10 years | 72 | 27.48 |
| More than 10 years | 147 | 56.11 |
| Citizenship status |  |  |
| Citizen | 199 | 75.95 |
| Non-citizen | 63 | 24.05 |
| **Recognition of specific conditions as a mental illness** | **n** | **%** |
| Identified "drug addiction" as a mental illness | 99 | 37.79 |
| Identified depression as a mental illness | 113 | 43.51 |
| Identified schizophrenia as a mental illness | 124 | 47.33 |
| Identified bipolar disorder as a mental illness | 133 | 50.76 |
| **Willingness to seek help from mental health professional** | **Mean (Standard Deviation)** | **P-value for mean difference** |
| For personal or emotional problems | 3.0 (SD=1.5) | 0.440 |
| For suicidal ideation | 3.1 (SD=1.5) |  |

**Table 2.** Primary and adjusted analyses.

| Knowledge about mental health and specific conditions | If you were having a <u>personal or emotional problem</u>, how likely is it that you would seek help from a mental health professional? |  | Predicted score 25% percentile of general knowledge | Predicted score 75% percentile of general knowledge |
| --- | --- | --- | --- | --- |
|  | *RR (95% CI)* | *P-value* |  |  |
| **General knowledge** |  |  |  |  |
| General knowledge | 1.06 (1.04, 1.07) | <0.001* | 2.50 | 3.45 |
| General knowledge, adjusted | 1.06 (1.04, 1.07) | <0.001* | 2.88 | 3.98 |
| **Specific knowledge** |  |  |  |  |
| Depression | 1.19 (1.06, 1.34) | 0.004* |  |  |
| Depression adjusted | 1.19 (1.06, 1.33) | 0.003* |  |  |
| Stress | 0.90 (0.79, 1.02) | 0.084 |  |  |
| Stress adjusted | 0.90 (0.79, 1.01) | 0.082 |  |  |
| Schizophrenia | 1.29 (1.15, 1.45) | <0.001* |  |  |
| Schizophrenia adjusted | 1.26 (1.12, 1.41) | <0.001* |  |  |
| Bipolar | 1.25 (1.12, 1.41) | 0.001* |  |  |
| Bipolar adjusted | 1.22 (1.09, 1.37) | 0.001* |  |  |
| Drug addiction | 1.26 (1.11, 1.42) | <0.001* |  |  |
| Drug addiction adjusted | 1.22 (1.08, 1.38) | 0.001* |  |  |
| Grief | 0.91 (0.81, 1.03) | 0.146 |  |  |
| Grief adjusted | 0.91 (0.81, 1.03) | 0.136 |  |  |
|  | **If you were experiencing suicidal thoughts, how likely is it that you would seek help from a mental health professional?** |  |  |  |
|  | **RR (95% CI)** | **P-value** |  |  |
| **General knowledge** |  |  |  |  |
| General knowledge | 1.05 (1.03, 1.06) | <0.001* | 2.57 | 3.38 |
| General knowledge, adjusted | 1.05 (1.03, 1.06) | <0.001* | 2.68 | 3.53 |
| **Specific knowledge** |  |  |  |  |
| Depression | 1.11 (0.98, 1.26) | 0.090 |  |  |
| Depression adjusted | 1.11 (0.99, 1.26) | 0.085 |  |  |
| Stress | 0.86 (0.76, 0.98) | 0.024* |  |  |
| Stress adjusted | 0.86 (0.76, 0.98) | 0.021* |  |  |
| Schizophrenia | 1.28 (1.13, 1.44) | <0.001* |  |  |
| Schizophrenia adjusted | 1.28 (1.14, 1.45) | <0.001* |  |  |
| Bipolar | 1.29 (1.15, 1.46) | <0.001* |  |  |
| Bipolar adjusted | 1.28 (1.14, 1.45) | <0.001* |  |  |
| Drug addiction | 1.21 (1.07, 1.37) | 0.003* |  |  |
| Drug addiction adjusted | 1.21 (1.07, 1.38) | 0.003* |  |  |
| Grief | 0.93 (0.82, 1.06) | 0.290 |  |  |
| Grief adjusted | 0.92 (0.81, 1.05) | 0.236 |  |  |

\* Each row represents the output from a single regression model. Adjusted models include age, education and ethnicity as covariates. Significant values $p > 0.05$ \*

### Primary Analysis

#### The association between general mental health knowledge and willingness to seek help from a mental health professional

After adjustment for age, education and ethnicity, a one-unit increase in mental health knowledge was associated with an increased probability of the average likelihood of seeking help from a mental health professional for personal or emotional problems (RR = 1.06 per point, CI: 1.04 – 1.07, p < 0.001). Stated differently, a study participant whose mental health knowledge was at the 25th percentile of the sample (14) had a predicted mean willingness level of 2.88 willingness to seek help from a mental health professional, whereas a study participant at the 75th percentile of mental health knowledge (20) had a mean of 3.98 willingness to seek help. Thus, an interquartile shift in mental health knowledge was associated with a 1.10 point difference in willingness to seek help, or a 38.19% change relative to the predicted mean willingness to seek help at the 25^th^ percentile of mental health knowledge. The estimated association between mental health knowledge and seeking help from a mental health professional for suicidal ideation was similar (RR = 1.06 per point, CI: 1.04 – 1.07, p < 0.001). Stated differently, a study participant whose mental health knowledge was at the 25th percentile of the sample (14) had a predicted mean level willingness of 2.68 to seek help from a mental health professional, whereas a study participant at the 75th percentile of mental health knowledge (20) had a mean of 3.53 willingness to seek help. Thus, an interquartile shift in mental health knowledge was associated with a 0.85 point difference in willingness to seek help, or a 31.72% change relative to the predicted mean willingness to seek help at the 25th percentile of mental health knowledge.

#### Recognition of specific mental illness conditions and its association with seeking help from mental health professionals (adjusted analysis)

After adjusting for age, education and ethnicity, those who endorsed that depression is a mental illness were more likely to report willingness to seek help from a mental health professional for personal or emotional problems, compared to those who did not endorse that depression is a mental illness (RR = 1.19, CI: 1.06 – 1.33, p = 0.003). Participants who recognized that schizophrenia, bipolar disorder, and drug addiction are mental illnesses were also more likely to report willingness to seek help for personal or emotional problems (RRs ranged from 1.22-1.26; all P<0.001). Participants who endorsed schizophrenia, bipolar disorder, and drug addiction are mental illnesses were more likely to report willingness to seek help for suicidal ideation (RRs ranged from 1.21-1.28; P-values ranged from 0.003 to <0.001). In contrast, respondents who accurately endorsed stress is not a mental illness had a decreased likelihood (14% less likely) to report willingness to seek help from a mental health professional for suicidal ideation (RR=0.86, CI: 0.76 – 0.98, p = 0.021).

#### Future Intended Stigma Behavior as an effect modifier

In our assessment of future stigma behavior as a moderator of the relationship between mental health knowledge and willingness to seek help, the interaction model did not yield statistically significant p-values (see table 3 in Appendix).

**Table 3.** Adjusted analysis showing effect modification of stigma behavior.

| Knowledge about mental health and specific conditions | If you were having a <u>personal or emotional problem</u>, how likely is it that you would seek help from a mental health professional? |  |  |  |  |  |
| --- | --- | --- | --- | --- | --- | --- |
|  | RR (95% CI) | P-value | Predicted score 25% percentile of general knowledge | Predicted score 75% percentile of general knowledge | RR for interaction (95% CI) | P-value |
|  | General knowledge |  |  |  |  |  |
| General Knowledge, high stigma | 1.06 (1.04, 1.09) | <0.001* | 2.74 | 3.95 | 1.02 (0.98, 1.05) | 0.327 |
| General Knowledge, low stigma | 1.05 (1.02, 1.07) | <0.001* | 3.05 | 3.99 |  |  |
|  | Specific knowledge |  |  |  |  |  |
| Depression, high stigma | 1.28 (1.08, 1.52) | 0.004* |  |  | 1.20 (0.94, 1.53) | 0.153 |
| Depression, low stigma | 1.07 (0.90, 1.28) | 0.446 |  |  |  |  |
| Stress, high stigma | 0.90 (0.74, 1.09) | 0.294 |  |  | 0.99 (0.77, 1.28) | 0.934 |
| Stress, low stigma | 0.91 (0.77, 1.08) | 0.268 |  |  |  |  |
| Schizophrenia, high stigma | 1.24 (1.04, 1.48) | 0.015* |  |  | 0.95 (0.75, 1.20) | 0.684 |
| Schizophrenia, low stigma | 1.30 (1.11, 1.53) | 0.001* |  |  |  |  |
| Bipolar, high stigma | 1.32 (1.12, 1.57) | 0.001* |  |  | 1.16 (0.92, 1.46) | 0.209 |
| Bipolar, low stigma | 1.14 (0.97, 1.34) | 0.101 |  |  |  |  |
| Drug addiction, high stigma | 1.33 (1.12, 1.57) | 0.001* |  |  | 1.13 (0.89, 1.44) | 0.312 |
| Drug addiction, low stigma | 1.18 (0.99, 1.43) | 0.071 |  |  |  |  |
| Grief, high stigma | 0.87 (0.71, 1.07) | 0.193 |  |  | 0.91 (0.70, 1.18) | 0.4703 |
| Grief, low stigma | 0.97 (0.82, 1.14) | 0.697 |  |  |  |  |
|  | If you were experiencing <u>suicidal thoughts</u>, how likely is it that you would seek help from a mental health professional? |  |  |  |  |  |
|  | RR (95% CI) | P-value | Predicted score 25% percentile of general | Predicted score 75% percentile of general |  |  |

|  |  |  | knowledge | knowledge |  |  |
| --- | --- | --- | --- | --- | --- | --- |
|  | General knowledge |  |  |  |  |  |
| General knowledge, high stigma | 1.05 (1.03, 1.08) | <0.001* | 2.14 | 2.89 | 1.01 (0.98, 1.05) | 0.559 |
| General knowledge, low stigma | 1.04 (1.01, 1.06) | 0.003* | 3.24 | 4.06 |  |  |
|  | Specific knowledge |  |  |  |  |  |
| Depression, high stigma | 1.13 (0.94, 1.35) | 0.205 |  |  | 1.07 (0.82, 1.40) | 0.604 |
| Depression, low stigma | 1.03 (0.86, 1.25) | 0.735 |  |  |  |  |
| Stress, high stigma | 0.89 (0.73, 1.09) | 0.273 |  |  | 1.00 (0.77, 1.31) | 0.983 |
| Stress, low stigma | 0.90 (0.75, 1.07) | 0.238 |  |  |  |  |
| Schizophrenia, high stigma | 1.20 (1.00, 1.45) | 0.052 |  |  | 0.83 (0.65, 1.06) | 0.134 |
| Schizophrenia, low stigma | 1.40 (1.19, 1.65) | <0.001* |  |  |  |  |
| Bipolar, high stigma | 1.32 (1.10, 1.57) | 0.002* |  |  | 1.01 (0.79, 1.29) | 0.931 |
| Bipolar, low stigma | 1.26 (1.06, 1.49) | 0.008* |  |  |  |  |
| Drug addiction, high stigma | 1.23 (1.03, 1.47) | 0.022* |  |  | 1.03 (0.79, 1.33) | 0.833 |
| Drug addiction, low stigma | 1.18 (0.97, 1.44) | 0.104 |  |  |  |  |
| Grief, high stigma | 0.94 (0.76, 1.17) | 0.603 |  |  | 1.01 (0.77, 1.34) | 0.918 |
| Grief, low stigma | 0.95 (0.80, 1.14) | 0.594 |  |  |  |  |

**Controlling for age, education and ethnicity. We report on moderation analysis based on high versus low stigma (future intended behavior). We show the RR for the interaction model between high versus low stigma. Significant p-values < 0.05\***

## Discussion

Our main finding was that there was a positive correlation between mental health knowledge and willingness to seek help from mental health professionals among Black adults after adjusting for age, education and ethnicity. Previous studies show that among Black adults, the conceptualization of mental illness has an influence on willingness to engage in help seeking behavior (1, 32). However, the specific association between mental health knowledge and use of mental health services among Black adults remains understudied (33, 34). The complexity of this relationship is exemplified in a study by Anglin and colleagues, in which the authors showed that Black people were more likely to believe that mental health professionals could help individuals with mental illness compared to white people but also more likely to believe mental illness would resolve without treatment (35), thus leading to delayed help seeking, disability and increased morbidity and mortality associated with poorly treated mental illness (1, 32, 36, 37). Notably, the perceived severity and knowledge of specific mental illnesses influenced anticipated treatment seeking behavior (35).

The specific recognition of mental illness is more strongly associated with greater willingness to use of mental health services compared to general knowledge about mental health. In particular, the specific knowledge of schizophrenia as a mental illness was associated with greater likelihood to be willing to use mental health services for personal and emotional problems. This is likely because knowledge about severe mental illness such as schizophrenia may be linked to an understanding of the need and value of professional mental health services (38), compared to knowledge about depression, commonly perceived to be less severe than schizophrenia (39). Previous studies show that people with schizophrenia are perceived as more dangerous and less likely to recover and this is even more pronounced for Black adults (40), whereas depression is seen more favorably in terms of severity and potential for recovery (39, 41). These perceptions may explain why in our study greater knowledge about schizophrenia was associated with greater use of mental health services for personal and emotional problems.

In our assessment of use of mental health services for suicidal ideation, the willingness to use mental health services was greatest for schizophrenia and bipolar disorder and lower for substance use disorder and no significant association was shown for depression. One hypothesis for these differences is twofold; on the one hand, in reference to specific knowledge about schizophrenia and bipolar disorder, the need for mental health services for suicidal ideation was greater than for personal or emotional problems. On the other hand, the perceived severity of schizophrenia and bipolar disorder as mental illnesses is greater than for depression or substance use disorder. Schizophrenia and bipolar disorder are known to carry an increased risk of suicide (8-30 times higher than the general population) and suicide attempts (42, 43), public campaigns present these illness as more severe than other mental illnesses (39).

While substance use disorder was also associated with willingness to use mental health services, for suicidal ideation, the likelihood to use services was lower for substance use disorder compared to schizophrenia and bipolar disorder. One explanation for this is that schizophrenia and bipolar disorder are more specific conditions than substance use disorder which includes several different illnesses that carry varying levels of perceived severity. Substance use disorder encompasses a wide variation of illnesses such as cocaine use disorder, opiate use disorder, cannabis use disorder and alcohol use disorder. Cocaine use disorder and opiate use disorder are perceived as more severe than alcohol or cannabis use disorder by the general public (44). Substance use disorder is often perceived as self-inflicted, hence there may be less perceived need for mental health services given the perceived role of will power. One study showed respondents held significantly more negative views of people with substance use disorder and were more skeptical of the effectiveness of mental health services for substance use disorder (45). Among Black adults, substance use disorder may be less associated with willingness to use mental health services because of the linkage between substance use disorder and criminalization of Black adults. Studies show that Black adults are disproportionately criminalized for substance use compared to white adults (46, 47).

In the association between knowledge of mental illness and willingness to use mental health services, we found high compared to low stigma did not have a moderating effect across knowledge areas. It is important to note that stigma reducing interventions target knowledge, attitudes and behavior. Given that stigma incorporates a dimension of knowledge (27, 28), it is possible that in our study because our measure of future intended stigma behavior does not incorporate specific knowledge as a component, we did not find stigma had a significant moderating effect on the primary association examined. The use of mental health services and its association with stigma is nuanced and requires further study, including a more nuanced understanding of knowledge as a component of stigma (for example, specific knowledge of the perceived illness severity and perceived course of specific mental illnesses).

Despite the severity of suicidal ideation, a strong risk factor for suicide attempt and suicide, there were no statistically significant mean differences between willingness to seek help for personal and emotional problems compared to seeking help for suicidal ideation. We expected a greater willingness to seek help for suicidal ideation (often a psychiatric emergency) across the independent variables of mental health knowledge but this was not consistently seen in our findings. There are low rates of mental health service use among Black adults (8, 9, 35), and some studies show lower likelihood to report suicidality to mental health providers due to medical mistrust among Black people (48-50). Therefore, based on our findings, building specific knowledge around mental health conditions, the specific conditions and spectrum of illnesses and the need for mental health service intervention for suicidality is critical among Black adults.

### Study limitations

This study was a cross sectional observational study, future studies would be needed to further assess longitudinal relationships between knowledge building and willingness to use mental health services over time. We present on 262 Black adults, while a modest sized study, previous studies that assess similar relationships across race and ethnicity tend to have low representation of Black adults with even smaller numbers than represented in this study (51, 52), we also present on a diverse representation of Black adults who identify as African Americans, African immigrants and Afro-Caribbean immigrants. We assessed one form of stigma behavior related to future intended stigmatization, it would be helpful to also analyze in future studies, other forms of stigma (self, experienced, anticipated, internalized or perceived) towards further assessing the moderating effect (53-58), if any, of stigmatization of mental illness. Our focus on one form of stigma related behavior in our assessment was because changing behavior is essential to improved engagement in care and we sought to reduce participant burden in the length of the questionnaire. Although, we did not measure actual behavior, we did measure future intended stigma behavior. Future studies would also benefit from a direct comparison between the effect of general knowledge and specific knowledge.

## Conclusion

These results have implications for how we build knowledge enhancing interventions that tend to focus on increasing knowledge about mental illness and optimizing engagement in mental health treatment in routine clinical care. Interventions that seek to increase specific knowledge may have better outcomes in improving utilization of mental health services by focusing on recognition of specific conditions, illness course, severity and risk of untreated mental illnesses. Given there are worse outcomes for Black adults around delayed treatment leading to increased chronic disability, higher rates of use of inpatient services due to severity at presentation and overall morbidity (5-9), our study highlights specific areas of knowledge that may serve as effective targets in construction of knowledge building interventions towards improved mental health outcomes for Black adults. In addition, while stigma behavior is important to address, we highlight the connection between knowledge and behavior related to use of mental health service may preclude stigma behavior as a barrier to mental health services. There are several implications highlighted for enhancing community education, public health awareness and optimization of engagement in mental health services for Black adults.

## Data Availability

All data produced in the present work are contained in the manuscript

## Appendix table

